# Temporal and Geographic Variation in Humidity as an Effect Modifier of the Association Between Heatwaves and Mental Health–Related Emergency Department Visits Among Medicaid Beneficiaries in the U.S. Gulf States

**DOI:** 10.64898/2026.11.20.26368609

**Authors:** Balaji Ramesh, David Luglio, Gregory A. Wellenius, Chowdhury Arafat Hossain, Md. Akib Al-Zubayer, Rica Pagay Monis, Md Mostafijur Rahman

## Abstract

With projected increases in humid heat, the extent to which humidity modifies the association between heatwaves and acute mental health (MH) outcomes is important but remains understudied. We evaluated this effect modification using Medicaid claims data on MH-related emergency department (ED) visits (2012-2019). The analysis included 14 million MH-related ED visits across five U.S. Gulf states, aggregated to county-weeks. Heatwave weeks were defined as weeks with daily maximum temperatures above the county-specific 97th percentile for ≥2 consecutive days. Following a county-level self-matched design, heatwave weeks were matched to non-heatwave weeks in the same calendar weeks of a randomly selected year. Conditional quasi-Poisson regression models estimated rate ratios (RRs) and 95% confidence intervals (CIs), with interactions assessing effect modification by weekly and county-level climatological humidity in separate models. Heatwave weeks were associated with higher rates of any MH-related ED visits (RR, 1.11; 95% CI, 1.09–1.12), including visits for substance use disorders, anxiety disorders, and depression. Associations were 7% and 5% stronger during high- and very-high-humidity weeks, respectively, than during low-humidity weeks indicating effect modification by humidity. These findings support incorporating humidity into heat-health interventions to better target populations and periods with heightened risk to reduce the MH burden associated with heatwaves.

## Introduction

Adverse mental health (MH) outcomes represent a substantial and growing public health burden in the United States and globally. MH conditions accounted for approximately 321 million disability-adjusted life years (DALYs), corresponding to 12% of the global disease burden, while in North America, they accounted for approximately 19.2 million DALYs, or 33% of the total disease burden ^1^. In the United States, the burden of MH conditions has increased substantially over the past decade ^2,3^. Concurrently, emergency department (ED) utilization for MH-related concerns has increased ^2,4,5^. For example, the proportion of ED visits with MH diagnoses increased by 65% between 2007 and 2016, with particularly two-fold increases among patients whose primary source of access was Medicaid ^5^. Over the years EDs have become an increasingly important source of primary and acute MH care, potentially due to limited access to affordable outpatient MH care, insurance carve-outs, and expanded Medicaid coverage under the Affordable Care Act ^2,4,5^.

There is strong evidence on the association between heat exposure and a wide range of adverse health outcomes, including MH outcomes ^6–9^. Physiological studies suggest that high humidity can exacerbate the adverse health effects of heat on human health ^10–13^. This may also apply to the MH impacts of heat, as studies suggest that at elevated temperatures cognitive tasks become more challenging under high-humidity than under low-humidity conditions ^14,15^, and hot and humid conditions may adversely affect sleep ^16,17^.

Epidemiological evidence on the joint effects of heat and humidity on mortality have been mixed^18–20^, while one study identified humidity as a strong effect modifier of cardiovascular morbidity ^21^. However, such epidemiological evidence on the modifying effect of humidity on heat-related MH outcomes remain limited. The only study examining this interaction in relation to psychosocial distress reported more than a two-fold increase in heat-related effects under high humidity, however its cross-sectional design and reliance on survey responses limited its ability to capture acute effects of heat ^22^. Other longitudinal studies of heat, humidity, and MH-related outpatient or ED visits did not assess effect modification, instead evaluated temperature and humidity as independent covariates or using composite temperature–humidity indices ^23,23–26^. Although composite indices are useful for heat-warning systems, they may obscure the independent contributions of temperature and humidity in evaluating epidemiological associations, highlighting the need to investigate effect modification by humidity ^10,18^.

Understanding this effect modification is particularly important given projections that climate change will intensify moist heat through increases in both air temperature and atmospheric moisture, particularly in tropical regions ^27^. Furthermore, understanding whether the association between temperature and MH outcomes varies across regions with different climatological humidity may help inform the development of region-specific interventions.

To address these research gaps, this study evaluated effect modification by specific humidity in the association between heatwaves and ED visits diagnosed with MH conditions using Medicaid claims data from 2012 to 2019 in the Gulf South region of the United States (Figure 1). We further investigated whether this association varied by county-level mean humidity, reflecting geographic differences in climatological humidity.

**Figure 1.**
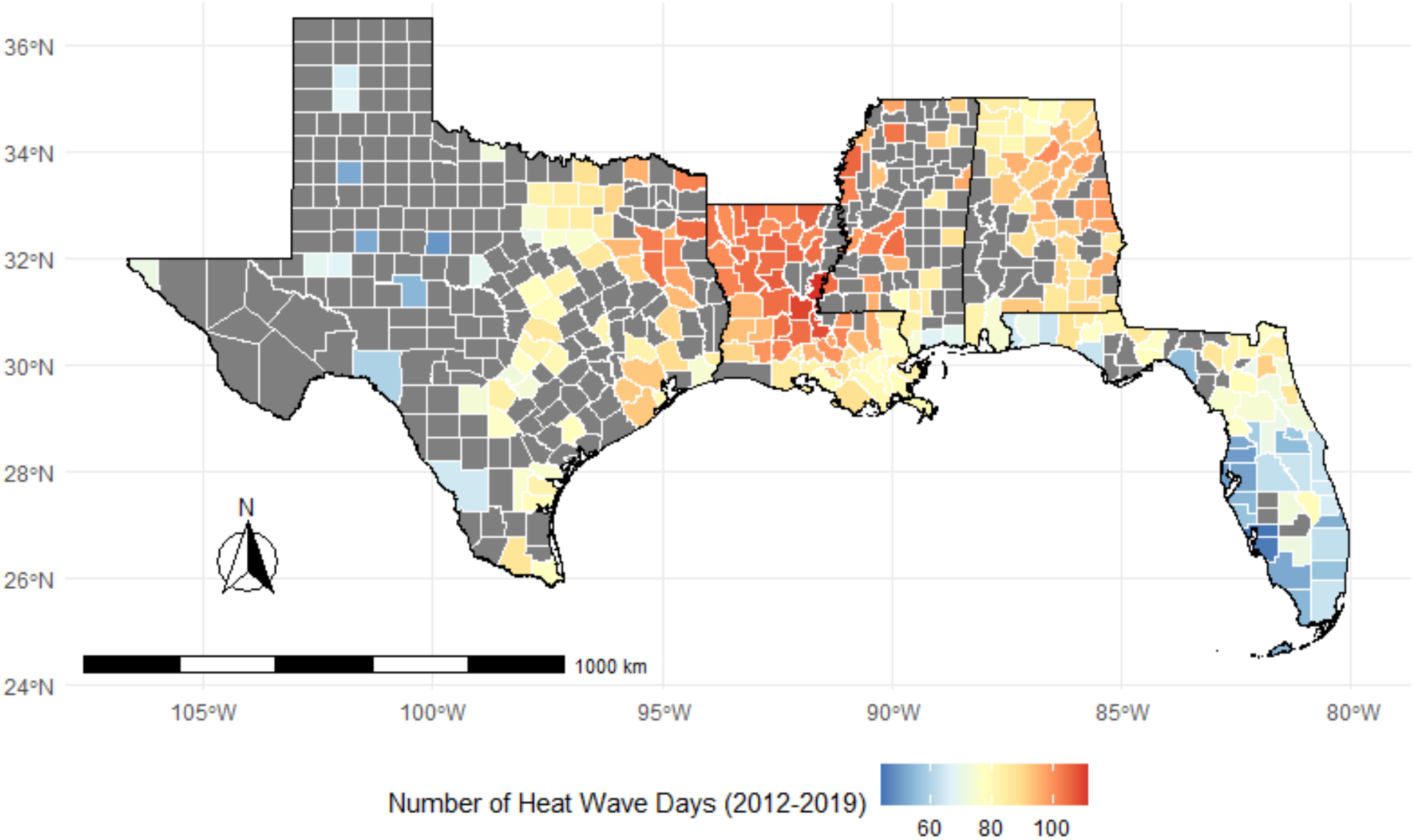
Number of heat-wave days by counties across the study area during 2012–2019. Gray-shaded were excluded due to insufficient data availability resulting from suppression of small counts in the Medicaid dataset.

The U.S. Gulf South warrants particular attention in this context, as it includes four of the five hottest states and four of the ten most humid states in the country and has also experienced increasing humidity over time ^28,29^. The region also faces substantial MH challenges, including higher prevalence of mental illness among youth, limited access to MH care, and higher rates of uninsured adults with mental illness ^30^. The focus on Medicaid beneficiaries further allow us to examine heat-related MH risks in a population that often experiences socioeconomic disadvantage, higher rates of chronic health conditions, and barriers to timely health care, and housing instability potentially increasing vulnerability to heat exposure ^31^.

## Methods

### Exposure data

Daily maximum ambient temperature and mean specific humidity data for 2012–2019 were obtained from the GridMET dataset for the five Gulf South states: Alabama, Florida, Louisiana, Mississippi, and Texas ^32^. The data were aggregated to the county level using population-weighted averages based on population estimates from the LandScan dataset ^33^. Heatwave days were defined as days on which ambient maximum daily temperature exceeded the 97th percentile maximum temperature for that county for at least two consecutive days.

Weekly mean specific humidity was calculated from daily values and were categorized into quartiles representing low (2.4–14.4 g/kg), moderate (14.5–16.3 g/kg), high (16.4–17.5 g/kg), and very high (17.6–20.9 g/kg) humidity weeks. Specific humidity (will be referred to as humidity hereafter) was preferred for this study than relative humidity because it provides a mass-based measure of atmospheric moisture with greater physiological relevance to sweat evaporation ^10^. To characterize geographic differences in climatological humidity, county-level mean humidity was calculated across the entire study period (2012-2019). Counties were then categorized into climatological humidity quartiles: low (8.35–13.3 g/kg), moderate (13.3–14.1 g/kg), high (14.2–15.4 g/kg), and very high (15.5–18.2 g/kg) humidity counties (Figure 2).

**Figure 2.**
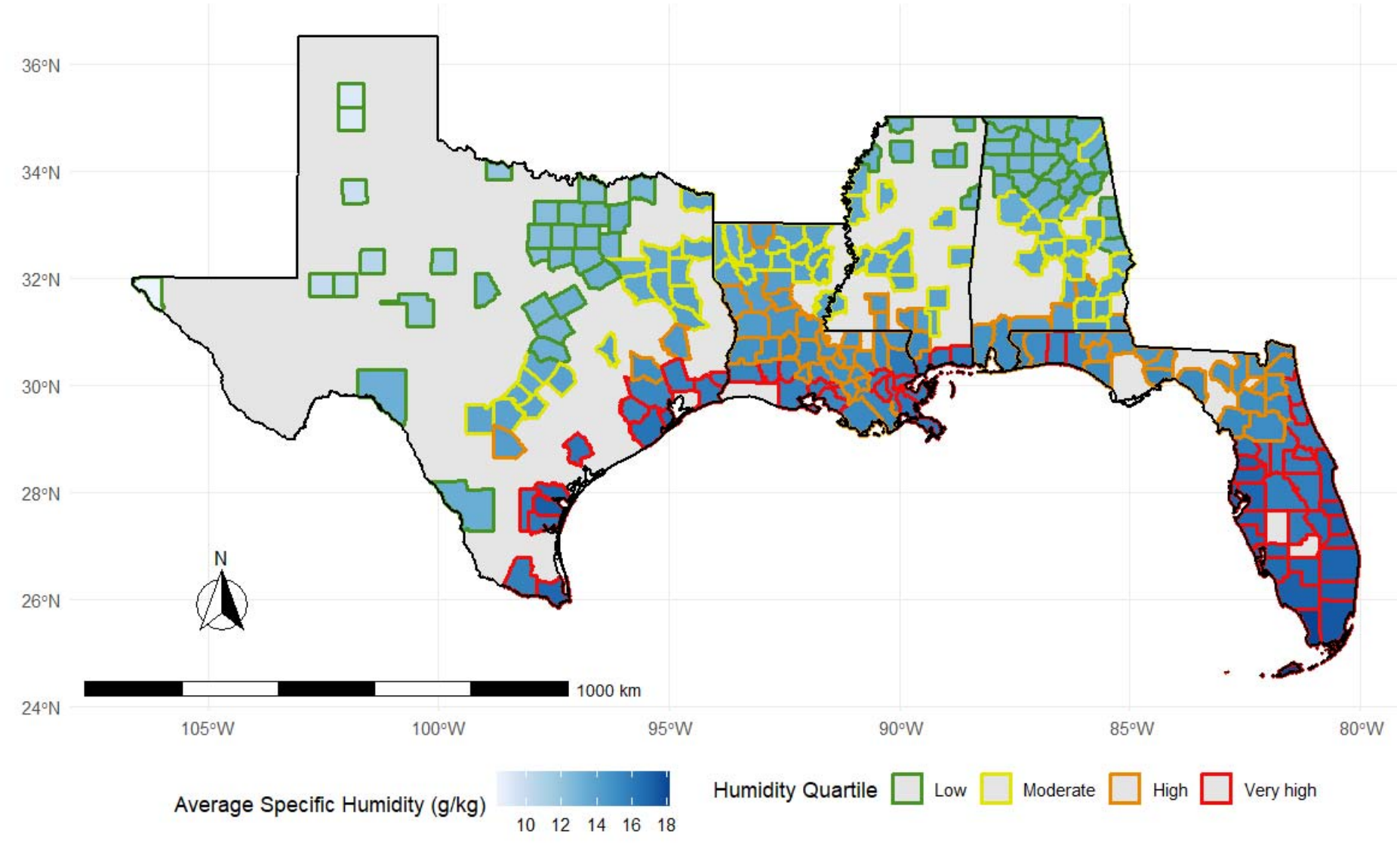
County-level climatological specific humidity, calculated as the mean specific humidity over the 2012–2019 study period. Blue shading represents mean specific humidity, while county border colors identify the corresponding humidity quartiles: Low, 8.35–13.3 g/kg; Moderate, 13.3–14.1 g/kg; High, 14.2–15.4 g/kg; and Very High, 15.5–18.2 g/kg

### Outcome Data

Weekly counts of ED visits diagnosed with MH conditions (hereafter referred to as MH-related ED visits) among Medicaid beneficiaries were obtained from Mathematica Inc. for the years 2012-2019 at the county level for the five states ^34^. These counts were derived using Medicaid claims data in the Medicaid Analytic eXtract (MAX) for years 2012–2015 and the Transformed Medicaid Statistical Information System (T-MSIS) Analytic Files (TAF) for 2014–202. The dataset included Medicaid beneficiaries aged 11 years or older, excluding those dually enrolled in Medicare and those aged 65 years or older. MH-related ED visits, including mental, behavioral, and neurodevelopmental disorders, suicidal ideation, suicide attempts, and intentional self-harm or poisoning, were identified using the corresponding the International Classification of Diseases, Ninth and Tenth Revision codes (ICD-9 and ICD-10) codes across all diagnosis fields in the claims (Supplementary Table 1) and counts were summarized across county-weeks. Small cell sizes (counts 1–10) were suppressed according to Centers for Medicare & Medicaid Services (CMS) data use agreements. The aggregated dataset comprised a total of 14 million ED visits.

Weekly county-level counts were also obtained for three specific categories of MH-related ED visits: anxiety disorders, depressive disorders, and substance use disorders (SUDs). These conditions were identified using their corresponding ICD-9 and ICD-10 codes (Supplementary Table 1). The study was approved by the Tulane University Institutional Review Board (IRB #2024-1441).

### Study design

To estimate the association between heat waves and MH-related ED visits at the weekly-county level, we adapted a county level self-matched study design. For each county, heatwave weeks were matched to the same calendar weeks in a randomly selected year in which no heatwave occurred during the index week or the preceding two weeks. These matched weeks will be further referred to as control weeks. This is similar to a case-crossover design but matches on exposure rather than outcome and has been used to investigate the health effects of environmental exposures ^35–38^

Further while matching heat wave to control weeks, as the data source transitioned from MAX to TAF during the study period, for each county matching was restricted to years within the same data source. This approach accounted for differences in diagnostic coding associated with the transition from ICD-9 to ICD-10, as well as potential differences arising from the transition between data systems. To minimize the potential impact of exposure misclassification, we excluded heatwave weeks in which the first heatwave day occurred after the fourth day of the week, along with their matched control weeks. Additionally, matched pairs in which neither the heatwave week nor the control week had any MH-related ED visits were excluded from the analysis.

### Statistical Analysis

Conditional quasi-Poisson regression models were used to account for overdispersion in the outcome counts while conditioning on the matched sets of heatwave and control weeks ^39^. The logarithm of the number of enrolled Medicaid beneficiaries in each country week was included as an offset to account for changes in beneficiary enrolment over time. Heatwave exposure was represented by a binary indicator distinguishing heatwave weeks from matched control weeks. The exponentiated coefficient of this variable, along with its 95% confidence interval (CI), represented the rate ratio (RR) for the association between heatwave exposure and MH-related ED visits. Associations were evaluated for the concurrent week (lag 0) and the following week (lag 1).

The within-county comparison achieved through the self-matching and conditional analysis controls for confounding by time-invariant county characteristics, while matching with same week-of-year accounts for seasonal patterns^39^. Long-term temporal trends were adjusted by including a natural spline of calendar year with 3 degrees of freedom. Weely average humidity was not treated as covariate due to its high correlation and to avoid imposing a hypothetical interpretation in which humidity remains constant while temperature changes, a scenario that is uncommon under ambient conditions ^10^. A sensitivity analysis additionally adjusted for humidity to assess the robustness of the estimated associations.

To assess the sensitivity of effect estimates to random control-week selection within the candidate years, we refitted the model using 1,000 independently generated sets of matched heatwave and control weeks. Relative risks (RRs) and corresponding 95% confidence intervals (CIs) were recorded across iterations to characterize variability attributable to the randomization in the matching procedure.

### Effect modification

Effect modification by humidity was evaluated by including an interaction between the binary heatwave exposure variable and humidity in models of any MH-related ED visits. Effect modification was assessed separately using county-level climatological humidity and weekly average humidity. Condition-specific outcomes (anxiety, depression, and SUD) were excluded from these analyses because sparse data limited the statistical power to estimate interaction effects.

County-level climatological humidity was categorized into quartiles. Thus, the interaction terms quantified the relative differences in the heatwave-associated RRs (on ratio scale) for counties in the moderate, high, and very high humidity quartiles compared with those in the low-humidity quartile.

Effect modification by weekly average humidity was evaluated using two approaches: (1) modeling humidity as a continuous variable and (2) categorizing humidity into quartiles, in which case the interaction terms estimated the relative differences in heatwave-associated RRs during moderate-, high-, and very-high-humidity weeks compared with low-humidity weeks.

All models were fitted using the gnm package in R ^40^. Humidity stratified estimates were extracted from the interaction terms using multcomp package in R ^41^.

## Results

Of the 524 counties in the study area, 248 contributed matched heatwave and control weeks to the final analytic dataset (Figure 1). The maximum number of Medicaid beneficiaries enrolled across these counties ranged from 3,740,950 in 2012 to 5,646,657 in 2019. In 2019, 23% of beneficiaries were non-Hispanic White, 25% were Black, 32% were categorized as other race/ethnicity, and 20% had unknown race/ethnicity. Females accounted for 59% of beneficiaries and males for 41%. By age group, 44% were aged 11–17 years, 21% were aged 18–26 years, 24% were aged 27–44 years, and 12% were aged 45–64 years.

The 97th percentile of maximum daily temperature during 2012–2019 across the study counties ranged from 33°C to 41°C (Supplementary Figure 1). A total of 2,723 county-weeks experienced heatwaves and were successfully matched to a control week. The crude rate of any MH-related ED visits was 241 per 100,000 beneficiary-weeks (95% CI, 236–245) during heatwave weeks, compared with 227 per 100,000 beneficiary-weeks (95% CI, 222–231) during control weeks. Overall, heatwave weeks had a 6% (95% CI, 3–9%) higher rate of any MH-related ED visits than control weeks.

### Association between heatwave weeks and MH-related Medicaid ED visits

Using the matched study designed and controlling for the long-term temporal trends, the conditional model estimated an RR of 1.11 (95% CI, 1.09–1.12) for any MH-related ED visits during heatwave weeks (lag0) compared with control weeks, indicating an 11% higher rate during heatwaves (Figure 3). Heatwaves were also associated with ED visits for substance use disorders (RR, 1.09; 95% CI, 1.07–1.11), anxiety disorders (RR, 1.06; 95% CI, 1.04–1.09), and depression (RR, 1.06; 95% CI, 1.03–1.10) (Figure 3).

**Figure 3.**
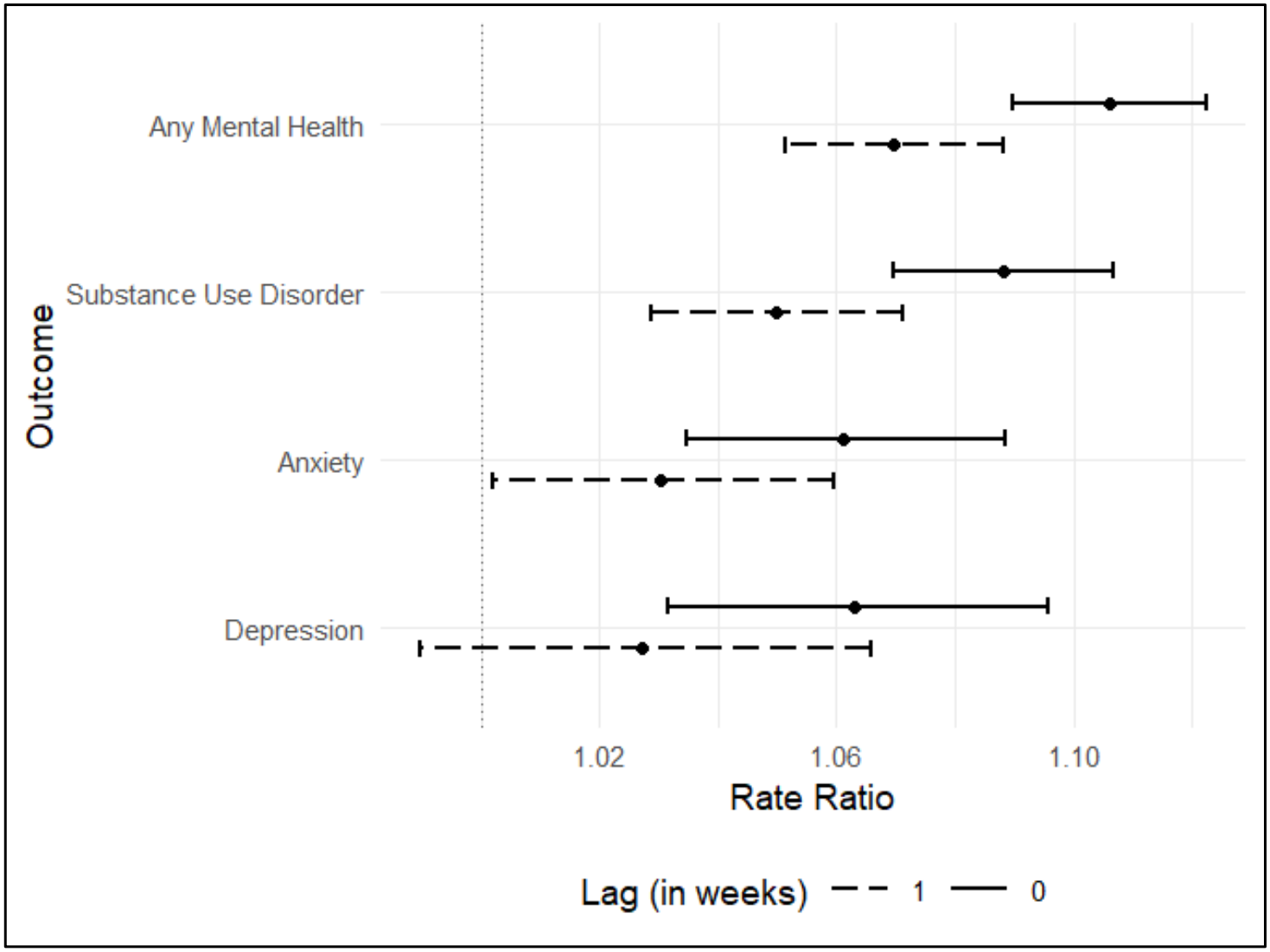
Rate ratios and 95% confidence intervals for the associations between heatwave and MH-related ED visits among Medicaid beneficiaries for the heatwave week (lag0) and the subsequent week (lag1).

Elevated rates of MH-related ED visits persisted into the week following the heatwave week (RR, 1.07; 95% CI, 1.05–1.09) when compared to the control weeks. This elevated rate was also reflected in SUD (RR, 1.05; 95% CI, 1.03–1.07) and anxiety related ED visits (RR, 1.03; 95% CI, 1.00–1.06). The lagged association with depression-related ED visits was more modest (RR, 1.03; 95% CI, 0.99–1.07) and did not reach statistical significance.

Sensitivity analysis adjusting for weekly average humidity showed similar results (Supplementary Figure 2). Across 1,000 iterations using independently generated sets of matched heatwave and control weeks, the estimated RRs ranged from 1.08 to 1.12 for any MH-related ED visits (Supplementary Figure 3). Results were consistent for anxiety and SUD-related ED visits, while the association with depression was statistically significant in 95.3% of iterations (Supplementary Figure 3). These findings indicate that the overall results were robust to random variation in control-week selection.

### Difference in association between heatwave and any MH-related ED visits by county-level climatological humidity

Among counties in the lowest quartile of climatological humidity, the RR for MH-related ED visits during the heatwave week was 1.07 (95% CI, 1.04–1.11) (Figure 4). The association was weaker among moderate humidity counties (RR, 1.03; 95% CI, 1.00–1.07) and stronger in high-humidity counties (RR, 1.14; 95% CI, 1.11–1.17) and very-high-humidity counties (RR, 1.12; 95% CI, 1.10–1.14) (Figure 4). Compared with low-humidity counties, the heatwave-associated RR was 6% higher in high-humidity counties (95% CI, 2%–11%; effect-modification *p* < 0.01) and 4% higher in very-high-humidity counties (95% CI, 0.3%–8% higher; effect-modification *p* = 0.03). The association in moderate-humidity counties did not differ significantly from that in low-humidity counties (*p* = 0.15).

**Figure 4.**
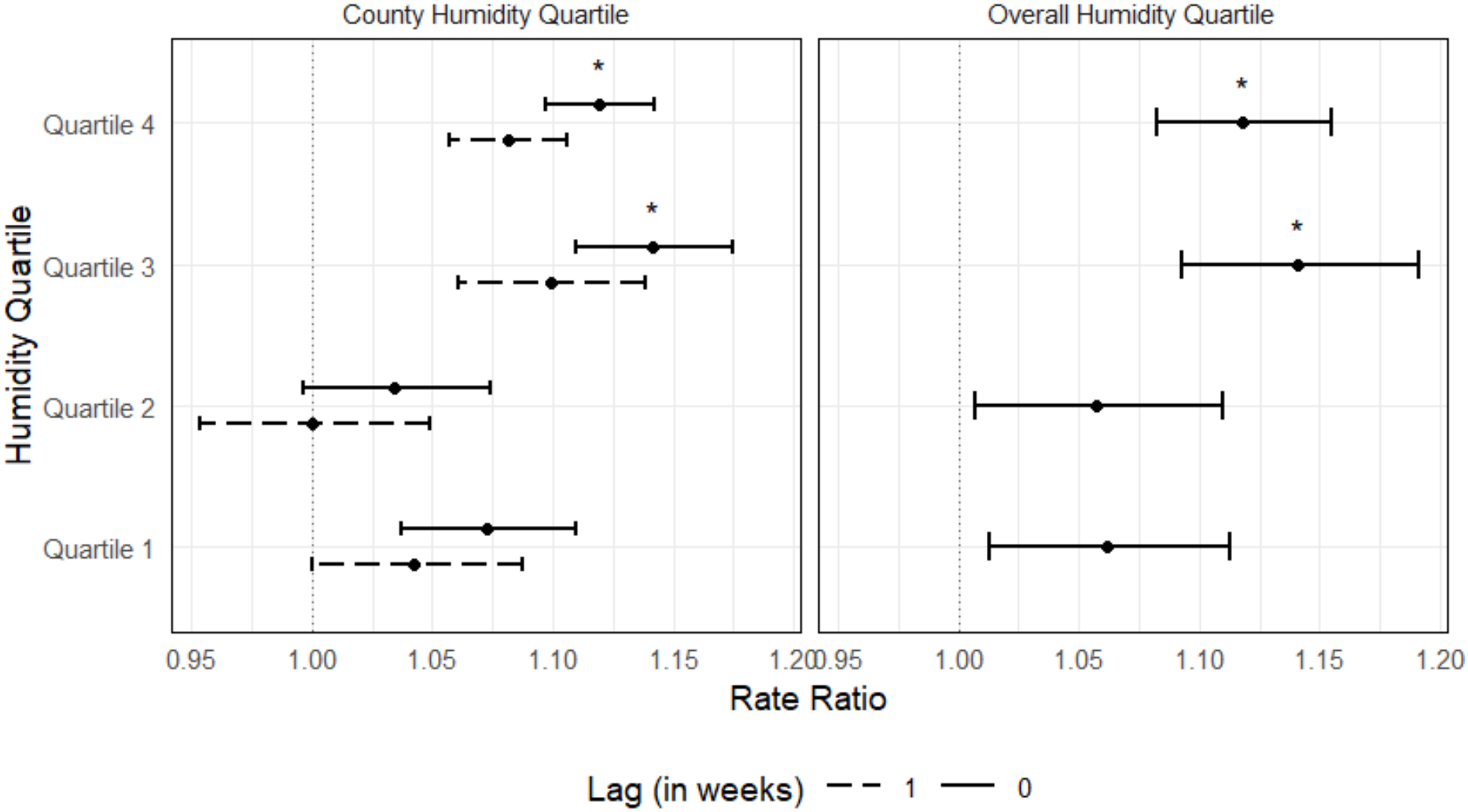
Rate ratios and 95% confidence intervals for the associations between heatwave and MH-related ED visits among Medicaid beneficiaries, stratified by humidity. The left panel presents estimates stratified by quartiles of county-level climatological humidity calculated over 2012–2019; the right panel presents estimates stratified by quartiles of weekly average humidity. * Indicates a statistically significant interaction effect compared with Quartile 1 (*p* < 0.05).

During the week following the heatwave week, elevated rates of MH-related ED visits were observed in high-humidity (RR, 1.10; 95% CI, 1.06–1.14) and very-high-humidity counties (RR, 1.08; 95% CI, 1.06–1.11). Weaker associations were observed in low-humidity (RR, 1.04; 95% CI, 1.00–1.09) and moderate-humidity counties (RR, 1.00; 95% CI, 0.95–1.05) (Figure 2). However, the RRs in high- and very-high-humidity counties did not differ significantly from those in low-humidity counties (effect-modification *p* = 0.06 and *p* = 0.13, respectively).

### Effect modification by weekly average humidity

Weekly average humidity distributions overlapped across counties classified into different climatological humidity quartiles (Supplementary Figure 4), indicating that counties with low climatological humidity also experienced weeks with relatively high humidity, and vice versa. This overlap indicates that county-level climatological humidity does not fully capture the variation in week-humidity conditions and supports evaluating effect modification by weekly average humidity.

During low-humidity weeks (2.4–14.4 g/kg), heatwave exposure was associated with a MH-related ED visits (RR, 1.06; 95% CI, 1.01–1.11). Similar associations were observed during moderate-humidity weeks (RR, 1.06; 95% CI, 1.01–1.11), whereas stronger associations were observed during high-humidity weeks (RR, 1.14; 95% CI, 1.09–1.19) and very-high-humidity weeks (RR, 1.12; 95% CI, 1.08–1.15) (Figure 4). Compared with low-humidity weeks, the heatwave-associated RR was similar during moderate-humidity weeks (effect-modification p = 0.88), and 7% higher during high-humidity weeks (95% CI, 2%–13% higher; effect-modification p < 0.01) and 5% higher during very-high-humidity weeks (95% CI, 1%–10% higher; effect-modification p = 0.02). Consistent with the quartile-based analysis, modeling weekly average humidity continuously and interacting with the binary heatwave indicator also indicated effect modification (*p* < 0.007).

## Discussion

This research shows that heatwaves that occurred during 2012–2019 were associated with increased weekly rates of MH-related ED visits among Medicaid beneficiaries, including ED visits related to depression, anxiety disorders, and SUD. These associations extended into the week following the heatwave, although the magnitude of the association was attenuated.

Further, weekly average specific humidity modified the association between heatwaves and MH-related ED visits, with higher humidity corresponding to a stronger association. One potential explanation for this pattern might be attributed to the role of humidity in thermoregulation. The 97th-percentile maximum temperature used to define heatwave days in our study counties was above 33°C, approaching the temperature range(>35°C) at which evaporative cooling (i.e., sweat evaporation) becomes increasingly important for dissipating body heat ^10^. However, as ambient humidity increases, the proportion of sweat that evaporates decreases, without contributing to evaporative cooling ^10,42^. This reduced evaporative cooling may increase physiological heat strain during heatwaves ^43^. Therefore, the stronger associations observed at higher levels of weekly humidity may partly reflect the heat strain experienced by the population under hot and humid conditions, potentially contributing to adverse MH outcomes that are observed in the ED visits.

Our findings on the heat-humidity interaction are consistent with findings from the cross-sectional study in Australia, which demonstrated that high ambient temperature and vapor pressure jointly increased psychological distress, with extreme humidity showing stronger effect modification for heat on psychological distress ^22^. Although other studies did not explicitly evaluate effect modification to compare with our findings directly, evidence from studies using composite temperature–humidity indices partly supports our findings. This includes increased psychiatric emergency medical service (EMS) dispatches and depression-related outpatient visits in China associated with high humidex levels, as well as an increased risk of self-reported depressive symptoms among survey respondents in India exposed to extreme wet-bulb heat stress ^24,44,45^. Similarly, a study in New York State found that the combined exposure to high temperature, high relative humidity, and high solar radiation was associated with the greatest excess risk of MH-related ED visits compared with the individual exposures ^23^.

The observed association between heatwaves and MH-related ED visits varied county-level climatological humidity. Counties in the high and very high humidity quartiles (third and fourth quartiles, respectively) exhibited stronger heatwave-associated increases in MH-related ED visits than counties in the lowest humidity quartile. These findings suggest that the adverse effects of heatwaves on MH-related ED visits may be greater in humid coastal regions, such as Houston, Texas; New Orleans, Louisiana; and Tampa, Florida, than in relatively drier inland regions, such as Dallas, Texas; Huntsville and Birmingham, Alabama; and Tupelo, Mississippi (Figure 2).

### Strengths

To our knowledge, this is the first longitudinal study to examine whether humidity modifies the association between heatwaves and acute MH outcomes. Muggy weather, characterized by hot and humid conditions, is widely recognized as uncomfortable, however their combined influence on MH in populations remains poorly understood. Our findings extend this evidence by demonstrating that the association between heatwaves and MH-related ED visits among Medicaid beneficiaries was stronger during periods of higher humidity.

The county-level self-matched study design strengthened our analysis by reducing potential bias from between-county confounding and strengthening causal inference. By examining both geographical variation in climatological humidity and temporal variation in weekly humidity, our study was able to demonstrate that humidity modified the association between heatwaves and MH-related ED visits across both spatial and temporal dimensions.

Our findings contribute to the growing body of evidence that extreme heat may exacerbate acute MH outcomes, particularly among the economically vulnerable Medicaid populations ^46,47^. Medicaid beneficiaries represent a population with a high proportion of individuals living in poverty and are disproportionately concentrated in the most socially vulnerable neighborhood ^48,49^. These low-income and socially vulnerable neighborhoods often have reduced access to heat-adaptation resources, such as air conditioning ^50^, and fewer protective environmental features, including green space ^51,52^, than the general population. These socioeconomic and environmental disadvantages may further amplify the MH burden of extreme heat as temperatures rise, underscoring the need for targeted interventions that promote equitable access to heat adaptation and protection.

### Limitations

This study has several limitations. First, exposure assessment at the county level may result in exposure misclassification because temperature and humidity can vary within counties. Such misclassification is likely to be nondifferential with respect to MH-related ED visits and therefore may have attenuated the observed associations toward the null. Second, the use of weekly data limits the ability to assess acute effects at daily lags and may have averaged out within-week peaks in humidity.

Third, we were not able to account for cross-lag interactions between heatwave and humidity (i.e., interactions between heatwave and humidity across different lag periods) as methods for estimating these effects remain underdeveloped and are not currently accommodated by commonly used distributed lag methods ^53^. Future methodological advances in this area may help address this limitation.

Fourth, our study was restricted to the Medicaid population and to counties with sufficient weekly ED visit counts to overcome small cell count suppression. These restrictions may limit the generalizability of our findings to the broader population of the Gulf South.

Fifth, this study focused on ED visits with any MH-related diagnosis (in primary or secondary diagnosis position) aggregated to county-weeks. This limited our ability to distinguish between different clinical circumstances underlying these MH-related ED visits. For example, we could not differentiate visits among patients with established chronic or subchronic MH conditions from those representing a first occurrence of an MH condition or an acute exacerbation of an underlying condition, such as an acute psychotic episode, manic episode, panic attack, or substance-use-related event. More detailed patient-level data would help disentangle how heatwaves may contribute to the onset or exacerbation of specific MH conditions.

Finally, the investigation of MH outcomes within the ED setting may not capture the overall burden of MH conditions in the population, particularly milder or less acute conditions for which individuals would seek care in other healthcare settings or may not seek care at all. For example, Ding et al. ^22^ found psychological distress was associated with both heat and humidity and, but these associations did not translate into increased treatment-seeking. Therefore, the observed associations in our study may represent only a portion of the broader effects of heat on MH. Examining MH outcomes across other healthcare settings would therefore be important for better understanding the broader extent of the effects of heat and humidity on MH.

## Conclusion

Heatwave were associated with higher weekly-rates of MH-related ED visits among Medicaid beneficiaries in the U.S. Gulf States, with stronger associations observed during weeks with high and very high humidity, indicating effect modification by humidity. Similarly, counties with higher climatological humidity exhibited stronger associations. These findings highlight the potential importance of considering humidity when identifying populations and periods of heightened vulnerability to heat-related MH impacts. Incorporating humidity into heat-health interventions, risk communication, and early warning systems may help inform strategies aimed at reducing the mental health burden associated with heatwaves.

## Data Availability

The Medicaid ED visit data are confidential and were purchased from Mathematica Inc. For the temperature and humidity data, please check the cited references.

## Acknowledgement

This research was supported by the Gulf Research Program of the National Academies of Sciences, Engineering, and Medicine [SCON-10001148; Rahman]. The authors declare no conflicts of interest.

## Code availability

The R codes used for the analysis are available at [Zenodo link]

