## Supplementary Information for "Temporal and Geographic Variation in Humidity as an Effect Modifier of the Association Between Heatwaves and Mental Health–Related Emergency Department Visits Among Medicaid Beneficiaries in the U.S. Gulf States"

### Supplementary Material

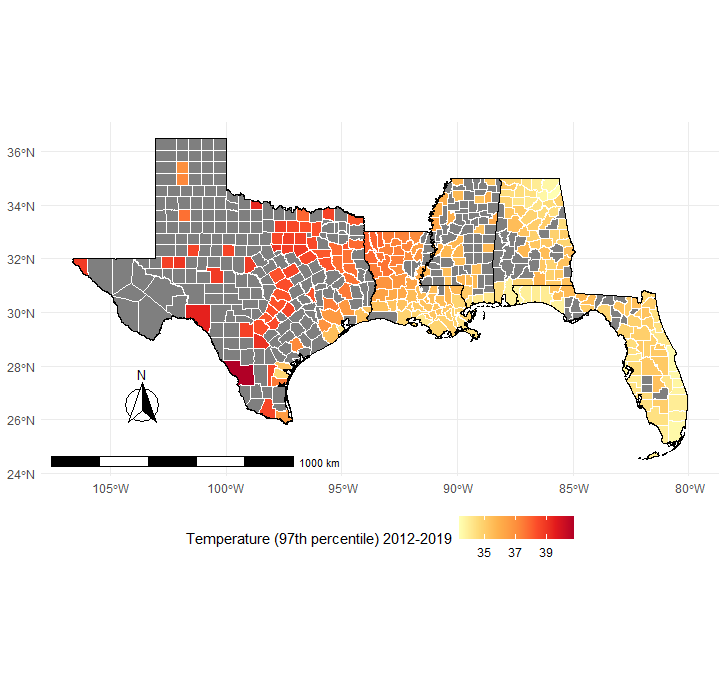

Supplementary Figure 1. County-level 97th-percentile maximum temperature across the study area during 2012–2019. Darker colors indicate higher temperature values. Black boundaries indicate state borders.

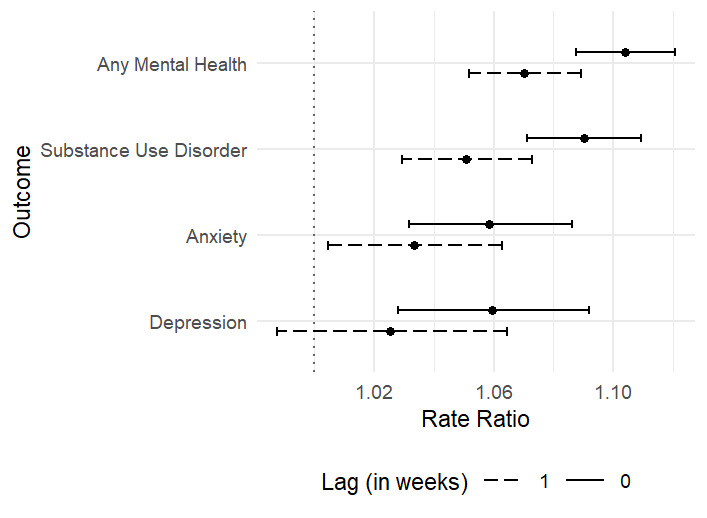

Supplementary Figure 2. Rate ratios and 95% confidence intervals for the associations between heatwave and MH-related ED visits among Medicaid beneficiaries for the heatwave week (lag0) and the subsequent week (lag1). Models were additionally adjusted for weekly mean specific humidity.

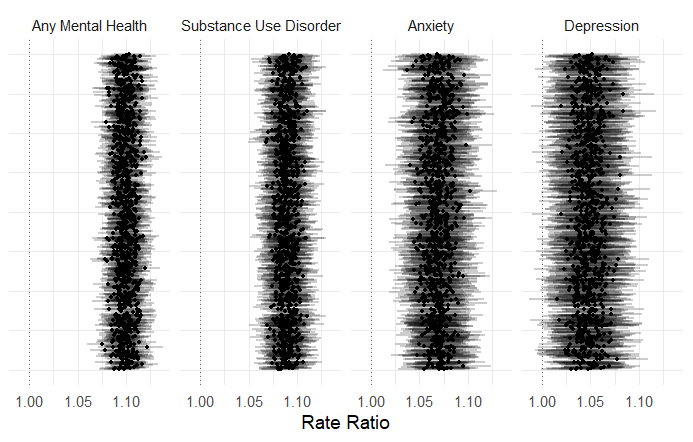
Supplementary Figure 3. Relative risks (RRs) and corresponding 95% confidence intervals (CIs) for the associations between heatwave and MH-related ED visits recorded across 1000 iterations to characterize variability attributable to the randomization in the matching procedure.

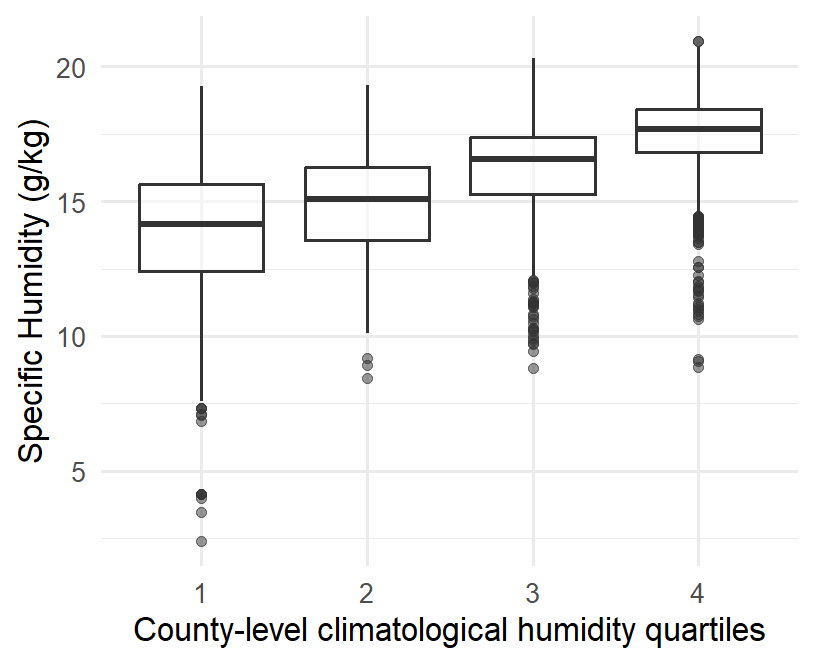

Supplementary Figure 4. Figure shows that even counties in the lowest county-average humidity quartile may have observations spanning multiple overall humidity quartiles, including relatively high-humidity values.

Supplementary Table 1. ICD 9 and ICD10 codes used for any mental health related and condition specific ED visits.

| Outcome | ICD-9 codes | ICD-10 codes |
| --- | --- | --- |
| All mental health | 290 – 319; E95.00 – E95.90; V62.84 | F01 – F99; T36 – T65; T14.91; X71 – X83; R45.851 |
| Anxiety | 293.84; 300.00; 300.01; 300.02; 300.09; 300.10; 300.20; 300.21; 300.22; 300.23; 300.29; 300.3; 300.5; 300.89; 300.9; 308.0; 308.1; 308.2; 308.3; 308.4; 308.9; 309.81; 313.0; 313.1; 313.21; 313.22; 313.3; 313.82; 313.83 | F06.4; F41.9; F41.0; F41.1; F41.8; F44.9; F40.9; F40.01; F40.02; F40.00; F40.10; F40.11; F40.210; F40.220; F40.218; F40.228; F40.230; F40.231; F40.232; F40.233; F40.240; F40.241; F40.242; F40.243; F40.248; F40.290; F40.291; F40.298; F40.8; F42; F48.8; F45.8; F48.9; F43.0; R45.7; F43.10; F43.12; F93.8 |
| Depression | 296.2x; 296.3x; 300.4; 301.12; 311 | F32x; F33x; F34.1; F39 |
| Substance use disorders | 291; 292; 303 – 305 | F10 – F19 |
